# Systematic Review of Studies on Telomere Lengths in Patients with Multiple Sclerosis

**DOI:** 10.1101/2020.11.23.20236992

**Authors:** Jan Bühring, Michael Hecker, Brit Fitzner, Uwe Klaus Zettl

## Abstract

**BACKGROUND:** Telomeres are protective cap structures at the end of chromosomes that are essential for maintening genomic stability. Accelerated telomere shortening is related to premature cellular senescence. Shortened telomere lengths (TL) have been implicated in the pathogenesis of various chronic immune-mediated and neurological diseases.

**OBJECTIVE:** We aimed to systematically review the current literature on the association of TL as a measure of biological age and multiple sclerosis (MS).

**METHODS:** A comprehensive literature search was conducted to identify original studies that presented data on TL in samples from MS patients. Quantitative and qualitative information was extracted from the articles to summarize and compare the studies.

**RESULTS:** A total of 51 articles were screened, and 7 of them were included in this review. In 6 studies, average TL were analyzed in peripheral blood cells, whereas in one study, bone marrow-derived cells were used. Four of the studies reported significantly shorter leukocyte TL in at least one MS subtype in comparison to healthy controls (*p*=0.003 in meta-analysis). Shorter telomeres in MS patients were found to be associated, independently of age, with greater disability, lower brain volume, increased relapse rate and more rapid conversion from relapsing to progressive MS. However, it remains unclear how telomere attrition in MS may be linked to oxidative stress, inflammation and age-related disease processes.

**CONCLUSIONS:** Despite few studies in this field, there is substantial evidence on the association of TL and MS. Variability in TL appears to reflect heterogeneity in clinical presentation and course. Further investigations in large and well-characterized cohorts are warranted. More detailed studies on TL of individual chromosomes in specific cell types may help to gain new insights into the pathomechanisms of MS.

**Highlights:**

- The relationship between aging and the pathophysiology and course of MS is not fully understood
- We have identified seven studies that analyzed telomere lengths (TL) in patients with MS
- Our meta-analysis revealed significantly shorter leukocyte TL in MS patients compared to healthy controls
- There is evidence that individual variability in biological aging reflects clinical heterogeneity in MS
- The potential use of TL as a biomarker of age-related disease mechanisms deserves further investigation

## 1. Introduction

Multiple sclerosis (MS) is an autoimmune disease, which affects the central nervous system (CNS). The disease usually begins in early adulthood [1], and approximately 2.8 million people live with MS worldwide [2]. MS is characterized by chronic inflammation, demyelination, gliosis and progressive axonal loss [3]. A hallmark of MS is the formation of focal lesions in the brain and spinal cord, which is mediated by the infiltration of immune cells [3]. The lesion sites are highly variable and thus every neurological symptom is possible. In consequence, the clinical presentation of individual MS patients is very heterogeneous, and symptoms can range from slight to very severe. Common initial symptoms of MS are optic neuritis, fatigue and paresthesia. The severity of MS is usually rated by the Expanded Disability Status Scale (EDSS), which considers cognitive and functional disabilities of MS patients and ranges from 0 (no abnormalities) to 10 (death as a result of MS) [4].

Most patients (85%) present with relapsing-remitting multiple sclerosis (RRMS), while 15% are diagnosed with primary progressive multiple sclerosis (PPMS) at disease onset [1,5]. RRMS is characterized by episodes of new or worsened preexisting symptoms (relapses), followed by periods of clinical stability (remission). In nearly 60% of RRMS cases, the disease turns after an average disease duration of 20 years into a secondary progressive course of MS (SPMS) [5]. Although patients with progressive MS typically do not experience relapses, they get steadily worse due to ongoing neurodegeneration, and superimposed attacks are possible [6].

The susceptibility to develop MS is up to three times higher for women compared to men [7,8], and female patients have an approximately 20% higher relapse rate [9]. Besides sex, former viral infections with Epstein-Barr virus [10,11] or human herpes virus 6A [12], vitamin D deficiency [10,13] and certain lifestyle factors, such as adolescent obesity and smoking [14], are established risk factors for MS [13,14]. Moreover, the individual risk of MS is determined by genetic predisposition, especially by genes of the major histocompatibility complex (MHC) [15]. Among the most prominent ones, HLA class II allele HLA-DRB1*15:01 increases the risk of MS [15-17], whereas class I HLA-A*02:01 has a protective effect [17]. Furthermore, about 200 non-MHC loci were found to be associated with MS susceptibility by international genome-wide association studies [18,19].

Within the last years, telomeres were shown to contribute to the pathomechanisms of various complex diseases [20,21]. Telomeres are nucleoprotein structures found at the end of each chromosome arm. In humans, they are composed of 5 – 15 kilobases (kb) long tandem repeats of the hexanucleotide TTAGGG [22,23] and six-subunit protein complexes, called shelterin [22,24]. The main roles of telomeres are the maintenance and protection of genetic information as well as the regulation of cellular replication capacity [24,25].

Telomere length (TL) shortens with age as a physiological process and subject to genetic and lifestyle factors (Figure 1). The length of telomeres decreases continuously because of the end-replication problem [22]. This refers to the issue that during every DNA replication cycle, the end of the lagging strand cannot be fully replicated. As a result, there is a loss of genomic sequence with every cell division. Accordingly, human telomeres shorten by approximately 50 to 70 base pairs per year [26]. In consequence, there is an upper limit in the number of cell divisions for a normal cell population, which is called the Hayflick limit [27]. Apart from aging, every process that provokes an accelerated shortening of TL can promote cellular senescence or apoptosis. In particular, telomeres are vulnerable to oxidative stress, which can be enhanced by smoking and dietary habits [22,28]. Moreover, chronic inflammation [29] and persistent viral infections [30,31] are well known to be associated with increased telomere attrition. In addition, lower physical activity was found to be linked with shorter telomeres [32].

**Figure 1:**
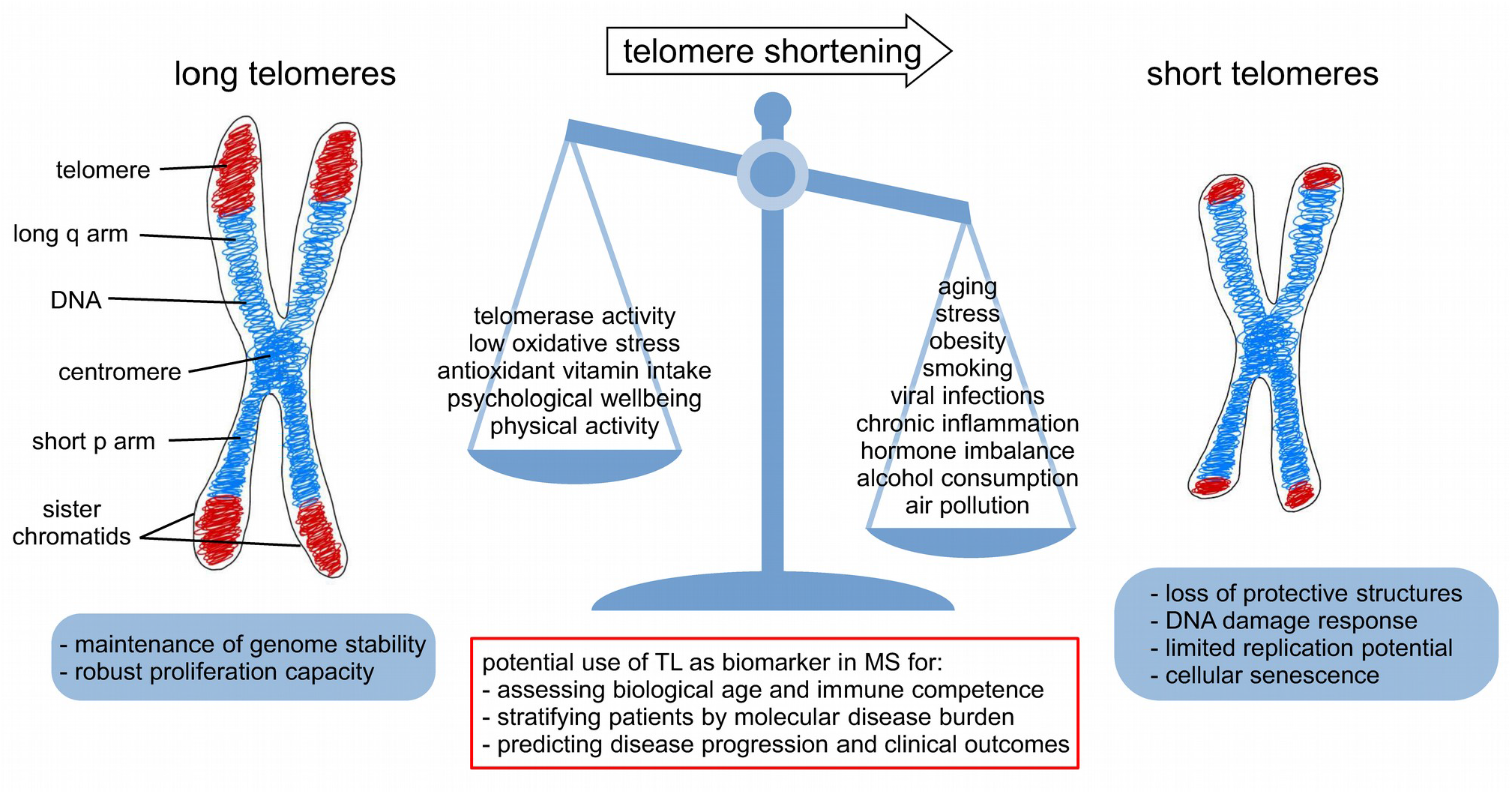
Factors contributing to accelerated telomere shortening. A metaphase chromosome is depicted on the left. The telomeres are highlighted in red and not drawn to scale in the diagram. They are located at the ends of the chromosome arms and consist of TTAGGG tandem repeat sequences (5 – 15 kb in humans). The centromere holds the two sister chromatids together. In normal cells, telomeres shorten with every cell division. Apart from individual genetic determinants that affect telomere length (TL), various environmental and lifestyle factors are known to influence telomere attrition. Disruption of telomere maintenance is associated with end-to-end chromosome fusion. Critically short telomeres trigger DNA damage responses such as cell cycle arrest. Telomere biology thus plays a crucial role in health and disease. In multiple sclerosis (MS), the analysis of TL may serve as a biomarker of the patients’ immunosenescence status to assess and predict clinical disease phenotypes.

Elongation and maintenance of telomeres is mainly controlled by a ribonucleoprotein complex called telomerase. It consists of the catalytic subunit telomerase reverse transcriptase (TERT) and the telomerase RNA component (TERC), which serves as a template for the addition of telomeric repeats. High telomerase levels are a hallmark of germline progenitor cells as well as embryonic stem cells, whereas most human cells display low or absent telomerase activity [33]. On the other hand, telomerase reactivation has been detected in 90% of all malignant tumors [34,35]. The transcription of TERT is regulated by many factors, including telomere length, hormones and viruses [22].

Diverse methods for the measurement of TL have been developed within the last 20 years. The traditional method is to assess the mean telomere restriction fragment (TRF) length of whole genomic DNA by Southern blot [36]. In 2002, Cawthon invented a real-time quantitative polymerase chain reaction (qPCR) assay [37]. In this method, a ratio of telomere signals (T) and single-copy gene signals (S), which are obtained in separate reaction wells, is calculated to determine relative T/S ratios that are proportional to the average TL of a biological sample [37]. Samples with a T/S > 1.0 have an average TL greater than that of the reference used. Cawthon later described an adapted monochrome multiplex qPCR (mmqPCR) assay, where the same well is used for amplification of both telomeric and single-copy DNA regions [38]. To measure telomeres of individual chromosomes, there are techniques such as quantitative fluorescence in situ hybridization (Q-FISH), universal single telomere length analysis (U-STELA) and telomere shortest length assay (TeSLA) for quantitation of the shortest telomeres [39], but each method has some limitations, as reviewed elsewhere [36,40].

Studies exploring TL as disease-associated biomarker typically use peripheral blood samples as source of DNA. Only few studies so far have investigated TL distributions in specific human cell subtypes and in non-blood disease-relevant tissues. However, recent reports suggest that whole blood TL is suited as a proxy for TL in many tissue types [41], even though the decline in TL with age varies between leukocyte subpopulations [42]. In comparison with healthy controls, shorter TL in leukocytes have been associated with several diseases including cardiovascular disease, type 2 diabetes, Alzheimer’s disease (AD) and autoimmune diseases, such as rheumatoid arthritis [20,21]. On the other hand, relatively long TL have been implicated in some types of cancer [43]. Since MS is a chronic inflammatory and neurodegenerative disease, telomeres may be linked to the development and course of this disease as well.

Here, we provide a comprehensive systematic review and meta-analysis of studies that analyzed TL in patients with MS. The studies were evaluated with regard to the respective study design and the employed TL measurement approach. We summarize the available evidence on the association of TL with MS subtypes. Moreover, we discuss how shorter TL may be involved in the molecular mechanisms underlying MS and whether telomeres as marker of cellular senescence may be suitable for predicting disease progression.

## 2. Methods

### 2.1. Search strategy and study selection criteria

For this review, we systematically searched for published articles on TL in MS patients. This was done by taking into account the Preferred Reporting Items for Systematic Reviews and Meta-Analyses (PRISMA) and by following the steps “identification”, “screening”, “eligibility” and “inclusion” [44]. Accordingly, we considered the current PRISMA recommendations and checklist in the design and conduct of this review.

The literature search was executed using the PubMed database as well as the preprint repositories bioRxiv and medRxiv to capture the latest research articles available. For identification of relevant papers, we wanted the search to be sensitive and potentially over-inclusive so that no relevant articles are missed. Therefore, only two terms, “multiple sclerosis” and “telomer*”, combined using the Boolean operator AND, were entered in the respective search interfaces. The asterisk wildcard symbol (*) was used for search term truncation. The latest check of this search was on November 19^th^, 2020. The reference lists of all included articles were also scanned with the aim to identify further studies on the same topic. The literature search was not limited by date of publication.

All articles were evaluated in full length in a standardized manner by two independent reviewers (JB and MH). The criteria for study selection were defined previously. We included studies that were written in English or German (I) and that appeared either in a scholarly journal or in an open access preprint repository. Moreover, we excluded papers that were no original research articles (II) and studies that did not include MS patients (III). Finally, we rejected studies that did not present data on TL (IV). Studies not meeting the inclusion criteria were excluded, and the reasons for exclusion were recorded. During the eligibility step, there was no restriction with regard to the course of MS, disease duration, disease severity, MS treatment, cohort size, race/ethnicity of patients and controls, type of sample material or TL measurement method used.

### 2.2. Data extraction

For comparing the remaining studies, relevant data was obtained from tables, graphs and statistical analyses described in the text. Data extraction was carried out using a predesigned data sheet by one reviewer (JB) and verified by another (MH). Any discrepancies were resolved by discussion. Accordingly, we created a table, which contained the main information for each study. More specifically, we collected information about the study design, the country of origin, the collection of biological samples (e.g., cross-sectional vs. longitudinal), the kind of cells that were used (e.g., peripheral blood leukocytes), the method utilized to determine TL (e.g., qPCR assay or TRF analysis) as well as clinical-demographic data about the patient cohort (e.g., the distribution of age and sex, the patients’ clinical course of MS [6] and the diagnostic criteria that were applied [45-47]). Additionally, we identified group comparisons (e.g., MS patients vs. controls, RRMS vs. PPMS, women vs. men) and strategies to control for confounding (e.g., matching or statistical regression) and recorded whether or not the findings reported in the respective studies reached statistical significance.

We have primarily conducted a narrative summary of the results, because the designs and methods of the included studies were quite different. Nonetheless, to further examine the relationship between TL and MS, a meta-analysis was performed as well. We used the metafor R package [48] for this analysis and for preparing a forest plot and a funnel plot. Means and standard deviations of TL that were reported in the studies for MS patients and controls were used to calculate standardized mean differences (SMD, also known as Cohen’s d) and 95% confidence intervals. A random-effects model was then fitted to the data to estimate the composite effect size. Heterogeneity across studies was evaluated by calculating the Q statistic *p*-value. Egger’s regression test was used to detect potential publication bias [49]. Due to the lack of data, we decided against additional subgroup analyses.

## 3. Results

### 3.1. Search results, study selection and overview

We ended up with 49 publications from our literature search in the PubMed database. Another 2 articles were found in bioRxiv and medRiv. No additional articles were found to be relevant by reference list screening. All articles were in English language (I). The full-texts of all articles were assessed for eligibility in accordance with the PRISMA statement guidelines [44]. The study selection process excluded most of the listed records as presented in the flow diagram (Figure 2): Five articles were excluded because they did not present original research (II). Subsequently, 18 studies were discarded because they did not contain an analysis of MS patient samples (III), and finally, 21 studies were rejected because they were not concerned with measuring TL (IV). In consequence, we included 7 studies in our systematic review [50-56] (Supplemental Table 1). Four of these studies also provided sufficient data to be pooled into a meta-analysis [51,52,55,56]. We then gathered diverse information from the selected articles, e.g., study design, sample size, patient characteristics, TL measurement method and cell population under scrutiny.

**Table 1:** General information about the 7 studies that were included in this review.

| First author (year) | Study design | Population | Diagnostic criteria | Sample size (patients : controls) | Disease course | Mean age in years (patients : controls) | % Female sex (patients : controls) | Matching |
| --- | --- | --- | --- | --- | --- | --- | --- | --- |
| Hug (2003) [50] | cases vs. controls, observational | Germany | Poser 1983 | 20 : 20 | RRMS: 20<br>SPMS: 0<br>PPMS: 0 | n.i. : n.i. | n.i. : n.i. | age |
| Guan (2015) [51] | cases vs. controls, observational | Asia | McDonald 2001 | 59 : 60 | RRMS: 19<br>SPMS: 20<br>PPMS: 20 | 33.3 : 34.3 | 45.8 : 43.3 | age, sex, lifestyle, diet |
| Guan (2018) [52] | cases vs. controls, interventional | Asia | McDonald 2001 | 34 : 44 | RRMS: n.i.<br>SPMS: n.i.<br>PPMS: n.i. | 32.8 : 33.6 | 47.1 : 47.7 | age, sex |
| Redondo (2018) [53] | cases vs. controls <sup>a</sup>, observational | United Kingdom | n.i. | 10 : 6 | RRMS: 0<br>SPMS: 5<br>PPMS: 5 | 54.1 : 55.7 | 30.0 : 33.3 | age, sex |
| Krysko (2019) [54] | case-only, observational | USA | McDonald 2001 | 516 : 0 | RRMS: 367<br>SPMS: 47<br>PPMS: 17<br>Other <sup>b</sup>: 85 | 42.6 : n.c. | 68.6 : n.c. | age, sex, disease duration, EDSS <sup>d</sup> |
| Habib (2020) [55] | cases vs. controls, observational | Germany | McDonald 2001 | 138 : 120 | RRMS: 102<br>SPMS: 27<br>PPMS: 5<br>Other <sup>c</sup>: 4 | 39.4 : 44.6 | 62.3 : 45.8 | - |
| Hecker (2020) [56] | cases vs. controls, observational | Germany | McDonald 2001/2005 | 60 : 60 | RRMS: 40<br>SPMS: 0<br>PPMS: 20 | 48.0 : 48.1 | 50.0 : 50.0 | age, sex |

In total, we have identified 7 studies in which telomere lengths (TL) were measured in patients with multiple sclerosis (MS). With one exception, all studies also included a control cohort. In the only interventional study, the impact of vitamin E supplementation on TL was explored. The patients were diagnosed according to the Poser criteria [45] or the (revised) McDonald criteria [46,47]. Furthermore, the study populations differed with regard to sample size, MS subtype composition, age distribution and proportion of women. The study cohorts were often matched for demographic characteristics to account for potential confounding in the TL analysis.
<sup>a</sup> = controls were not healthy (controls underwent total hip replacement for indication of osteoarthritis)
<sup>b</sup> = comprises 80 patients with clinically isolated syndrome, 4 patients with progressive relapsing MS and 1 patient with an unclear diagnosis
<sup>c</sup> = subtype of MS unclear
<sup>d</sup> = matching was assured for a subset of SPMS converters and non-converters (n=46) with TL measured longitudinally (no control cohort in this study)
EDSS = Expanded Disability Status Scale; n.c. = no control group; n.i. = no information; PPMS = primary progressive MS; RRMS = relapsing-remitting MS; SPMS = secondary progressive MS

**Figure 2:**
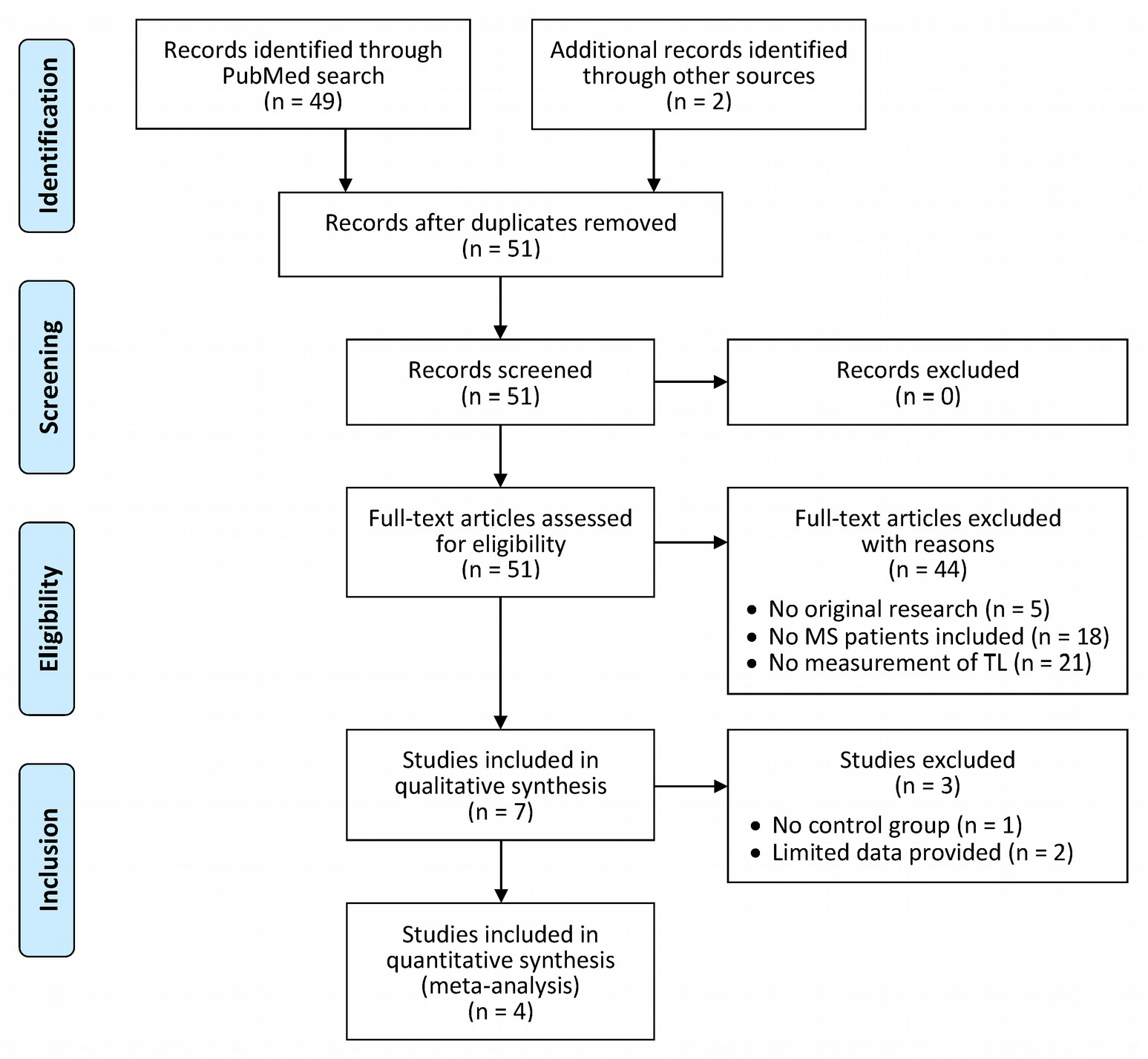
PRISMA flow diagram of the study selection process. Following the Preferred Reporting Items for Systematic Reviews and Meta-Analyses (PRISMA statement) [44], we considered the four steps identification, screening, eligibility and inclusion to identify studies for our systematic review. The boxes that are connected through arrows show the steps of the selection process and the number of articles that were included or excluded. A total of 51 articles were found in the literature databases and evaluated for eligibility. The study selection finally revealed 7 studies on telomere lengths (TL) in patients with multiple sclerosis (MS). A subset of 4 studies was included in the meta-analysis.

The seven included studies were published within the last five years, except the study by Hug et al., which was already published in 2003 [50]. The studies were conducted by six different research groups with either Asian (n=2), European (n=4) or American (n=1) MS patient cohorts. In all studies, the collection of patient samples was performed in a cross-sectional manner, while three studies additionally used longitudinal blood sample series obtained over a period of up to >10 years [52,54,56]. With one exception [54], all studies compared TL between MS patients and controls. The number of patients included in the investigation of telomeres ranged from 10 [53] to 516 [54]. In the following section, we have summarized the objectives and findings for each of the individual studies.

### 3.2. Characteristics and results of the included studies

The first study was published by Hug et al. [50]. They explored whether there are TL differences in T cells between RRMS patients (n=20) and age-matched healthy controls (n=20). For this purpose, mean TRF lengths were determined for CD4+ and CD8+ cells that were isolated by magnetic separation (positive selection) from peripheral blood mononuclear cells. The research group observed a decline of TL with age in MS patients as well as in controls in both cell populations. However, they did not find any statistically significant difference in relative TL or age-dependent telomere shortening between the patients and controls. The TL were also similar in the two T cell subsets: The CD4+ and CD8+ T cells had a mean TL of 10.1 kb and 9.2 kb in patients with MS and 8.8 kb and 9.4 kb in controls, respectively. Hug et al. also investigated the enzymatic activity of telomerase in the circulating T cells. Telomerase activity was detectable at low levels and was slightly higher in CD4+ cells compared with CD8+ cells, but there was no significant difference between the MS patient group and the healthy control group. The authors concluded that there is no evidence of an accelerated T cell turnover, but that an altered T cell composition in MS may interfere with peripheral immune tolerance mechanisms.

The study by Guan et al. from 2015 [51] enrolled 59 patients with MS (benign RRMS: n=19, SPMS: n=20, PPMS: n=20) and 60 healthy individuals. The MS and control groups were similar with regard to age and sex distribution as well as lifestyle and dietary habits. Subjects with infections, diabetes, habitual smoking status or corticosteroid use within the past 3 months were excluded. The average TL in peripheral blood cells was then assessed by TRF analysis. The authors found significantly shorter TL, on average, in PPMS patients (6.5 kb in men and 7.0 kb in women) compared to controls (8.4 kb in men and 10.1 kb in women), whereas the TL in RRMS and SPMS patients did not differ significantly from those in controls. Guan et al. also reported a clear negative association of TL and age for the control group, regardless of sex, and for male SPMS patients. Moreover, they evaluated markers of oxidative stress in plasma and urine. This revealed significantly elevated levels of urinary 8-iso-PGF2α (a marker of lipid peroxidation) and reduced antioxidant capacity of plasma lipoproteins in MS, especially in PPMS. However, there was no significant relationship between TRF results and oxidative stress markers in either group of subjects. In sum, these data suggested that enhanced oxidative damage and shorter somatic TL distinguish in particular MS patients with a primary progressive course of disease.

The subsequent study by Guan et al. from 2018 [52] analyzed TL and systemic peroxidation in 34 MS patients and 44 healthy subjects. The two cohorts had a similar female:male ratio and average age, and the exclusion criteria were as in the former study [51]. However, in this study, MS patient samples were collected at two time points to evaluate the biological and clinical effects of antioxidant vitamin E supplementation. Accordingly, half of the patients received α-tocopherol (400 mg/day) for 3 months and the other half (matched for age and sex) did not. TL were measured by TRF analysis using genomic DNA from peripheral blood (i.e., circulating leukocytes). Additionally, Guan et al. employed serum samples for testing oxidation of low-density lipoproteins (LDL) and urine samples for determining 8-iso-PGF2α concentrations. As a result, in line with their previous study, significantly shorter average TL were observed in MS patients (6.9 ± 1.0 kb) compared to controls (9.1 ± 1.4 kb). The MS group was also characterized by significantly higher levels of urinary 8-iso-PGF2α and lower antioxidant capacity in serum. Following vitamin E administration, 8-iso-PGF2α levels were significantly reduced by nearly 20%. However, the treatment over 3 months appeared not to be long enough to detect significant changes in TL or clinical parameters (e.g., EDSS scores). This study thus confirmed shorter TL in MS patients and indicated that antioxidants, such as vitamin E, may be useful to attenuate enhanced oxidation conditions present in MS.

Redondo et al. [53] used bone marrow-derived mesenchymal stromal cells (MSC) from 10 patients with progressive MS and 6 controls with osteoarthritis. These multipotent MSCs were expanded in vitro and harvested at passage number P2 and P6 to measure TL by TRF analysis. The authors found no significant difference in TL between samples derived from controls and MS patients. Duration of disease progression and subtype of progressive MS were also not associated with TL. However, significant effects of age (*p* = 0.024) and passage number (*p* < 0.0001) were detected by multivariable regression: TL correlated negatively with age and decreased from P2 to P6 in both control and MS MSCs. This decrease only reached statistical significance in MS MSCs. Moreover, these cells were characterized by reduced in vitro expansion potential. Redondo et al. thus concluded that telomere shortening is accelerated in MSCs from MS patients, suggesting that premature cellular aging and senescence may contribute to the pathophysiology of MS.

Krysko et al. [54] performed the so far largest and most elaborated study on TL in MS. They used whole blood DNA samples of 516 patients, including 80 individuals with clinically isolated syndrome (CIS) and 367 RRMS patients. These patients were studied for up to 10 years from baseline by clinical assessments and magnetic resonance imaging (MRI). For a subset of 46 individuals, DNA samples from multiple time points were analyzed. This subset consisted of a group of patients who converted to SPMS (n=23) and a group of patients who remained in the RRMS stage (n=23), while matching for potential confounders such as age and sex. The average leukocyte telomere lengths (LTL) were then measured by real-time qPCR [37], and the resulting T/S ratios were scaled by −0.2 to ease the interpretation of regression coefficients. The cross-sectional analyses over the entire patient cohort revealed that higher age and disease duration were clearly associated with shorter LTL (*p* < 0.001), while there was no significant association between LTL and sex, body mass index (BMI), smoking status or type of MS treatment. More importantly, shorter telomeres were associated with greater disability and brain atrophy, after adjusting for age, sex and disease duration: For every 0.2 unit lower LTL, the baseline EDSS score was 0.27 units higher (*p* < 0.001) and total brain volume and white matter volume were 7.4 mm^3^ and 4.0 mm^3^ lower, respectively (*p* < 0.05). The effect of age on EDSS that is mediated by LTL was estimated to be 15.1% by mediation analysis. The authors also assessed the association of baseline LTL with change in clinical and MRI outcomes over time. Based on adjusted models, they found LTL to be weakly associated with the rate of change in EDSS (*p* = 0.06) and cortical gray matter volume decline (*p* = 0.02). Moreover, among the patients with RRMS, for every 0.2 unit decrease in LTL, a 1.27 times higher relapse rate was noted (*p* = 0.001). In the subset of 46 patients with LTL measured longitudinally over up to 10 years of follow-up, a −0.2 unit change in LTL was associated with an EDSS score increase by 0.34 (*p* = 0.012). In this subset, patients also had 1.4 times the odds of converting from RRMS to SPMS for every 0.2 unit lower baseline LTL, although this finding did not reach statistical significance (*p* = 0.40). However, it has not been reported whether there were also differences in average LTL between patients with different subtypes of MS in the full cohort. In sum, this study convincingly showed that TL is associated with disability progression independent of age and disease duration, suggesting that individual variability in biological aging may contribute to clinical heterogeneity in MS.

Habib et al. [55] assessed TL in whole blood DNA samples of 138 MS patients and 120 healthy controls. Three MS groups were distinguished: RRMS (n=102), SPMS (n=27) and PPMS (n=5), while 4 patients had an undefined course of MS. The RRMS patients were the youngest (average age: 35.2 years), followed by healthy controls (44.6 years), PPMS patients (49.0 years) and SPMS patients (51.8 years) (*p* < 0.001). The groups were also not matched by sex (MS: 37.7% male, controls: 54.2% male). Relative LTL were determined using the mmqPCR method by Cawthon [38]. Significantly shorter LTL were observed in all 3 groups of MS patients (mean T/S: RRMS: 0.76 ± 0.22; SPMS 0.67 ± 0.21; PPMS 0.67 ± 0.27) as compared with the controls (0.94 ± 0.25) (*p* < 0.001 with adjustment for age as covariate). No significant difference in TL was observed between men and women, but a strong negative relationship between age and TL was seen in both controls and MS patients (*p* < 0.001). Accordingly, the patients with a progressive course of disease (PPMS and SPMS), which were much older, had shorter telomeres than the patients with RRMS. Shorter TL also correlated with higher EDSS scores (*p* = 0.001), but this association was again driven by age. Additionally, Habib et al. conducted an in vitro experiment using lymphoblastoid cell lines (LCL) generated from peripheral blood B cells. This analysis revealed shorter TL in LCL from MS patients (n=3) compared to controls (n=3) but overall similar dynamics of telomere loss over up to 15 passages of cultivation in both groups. The authors therefore speculated whether accelerated telomere shortening may precede MS onset.

The most recent study by Hecker et al. [56] comprised 40 RRMS patients, 20 PPMS patients and 60 healthy controls. Those three groups were well matched for age (with an average of 48.0 years and 48.1 years for patients and controls, respectively) and sex (female:male ratio always 1:1). The average LTL was determined for all subjects by multiplex qPCR [38] using DNA from peripheral whole blood samples. The data were analyzed in relation to the long-term clinical course of the patients. To this end, relapses and progression of disability were evaluated over a period of up to 10 years after the blood sampling. For a subset of 10 MS patients, a second blood sample was collected after this follow-up period. In the longitudinal analysis, LTL decrease over time was seen for all patients (*p* < 0.001), independent of disease subtype. In the cross-sectional analysis, significantly shorter telomeres could be detected in the group of RRMS patients (mean T/S = 0.92) compared to the PPMS patients (mean T/S = 1.16) and controls (mean T/S = 1.12) (*p* = 0.003). No significant differences were found between the PPMS group and controls as well as between women and men. However, age was negatively associated with LTL in all 3 study cohorts. Hecker et al. divided the subjects into two groups with relatively short (T/S ratio <1) or long telomeres (T/S ratio >1). Patients with RRMS were less likely to belong to the long telomere group (odds ratio = 0.399, *p* = 0.032), and those with relatively short LTL were also at much higher risk to convert to SPMS in the 10-year follow-up (hazard ratio = 8.308, *p* = 0.050) in age-adjusted tests. On the other hand, the LTL of RRMS patients at baseline had little value in terms of predicting the rate of change in EDSS and the risk of subsequent relapses. Nevertheless, these data suggest that LTL as a biomarker for immunosenescence may capture age-related disease mechanisms.

The following section presents an integrative comparison of study methods and results that were presented in the papers included in this review.

### 3.3. Comparative assessment of studies on TL in MS

Five of the identified studies included a healthy control group (Table 1), while Redondo et al. [53] used samples of patients with osteoarthritis as controls. The study by Krysko et al. [54] is the only one that did not include a control group. However, their study comprised the largest number of MS patients (n=516) with the aim to detect associations of LTL with changes in clinical and MRI metrics over a follow-up period of 10 years. Differences between the studies were also noted with regard to the MS population under scrutiny (Table 1): In the majority of studies, the diagnosis of MS was confirmed using the original or revised McDonald criteria from 2001/2005 [46,47]. The early study by Hug et al. [50] was based on patients diagnosed with relapsing MS according to the Poser criteria from 1983 [45], whereas Redondo et al. [53] did not specify the applied diagnostic criteria and considered only cases with progressive MS. Differences in LTL between subgroups of MS patients with different courses of disease (RRMS, SPMS and PPMS) were examined in 3 of the 7 studies [51,55,56]. Patients with CIS were included only in the analysis by Krysko et al. [54]. The second study by Guan et al. [52] is currently the only one that investigated the effects of a therapeutic intervention (vitamin E administration) on TL in blood cells of MS patients.

More men than women were included in the studies by Redondo et al. [53] and Guan et al. [51,52], whereas the proportion of female MS patients was 68.6% in the study by Krysko et al. [54]. There were also huge differences between the studies with regard to the age of the patients: For instance, the patients in the studies by Guan et al. had an average age of 32.8 years [52] and 33.3 years [51], respectively, whereas the TL analysis by Redondo et al. was conducted in a much older group of MS patients, with a mean age of 54.1 years [53]. Notably, the latter two research groups as well as Hecker et al. [56] paid specific attention to ensure that the cohorts to be compared were well matched in terms of age and sex. There was no control group in the study of Krysko et al., but in the subset of patients that had LTL measured over time for comparing SPMS converters (n=23) and non-converters (n=23), a careful matching for age, sex, disease duration and EDSS score was carried out [54]. On the other hand, the cohorts in the study by Habib et al. had uneven age and sex distributions [55], and Hug et al. only reported that the MS patients and healthy controls were age-matched [50].

In 5 out of the 7 reviewed studies, whole blood samples were used for TL analysis (Table 2). In these studies, leukocytes are the main source of telomeric DNA as human erythrocytes and platelets have no cell nucleus. The measurement of leukocyte TL provides a rough insight into the average length of telomeric ends in a heterogeneous population of granulocytes, lymphocytes and monocytes. So far, only the study by Hug et al. [50] used specific immune cells, namely peripheral blood CD4+ and CD8+ T cells, for assessing TL in MS patients. As an exception, Redondo et al. [53] made use of MSCs that were isolated from bone marrow samples, as they were interested in exploring the bone marrow microenvironment in the context of autologous MSC-based therapy for progressive MS. In their study as well as in the study by Habib et al. [55], experiments with patient-derived cells maintained in culture were performed to examine the dynamics of telomere attrition in vitro.

**Table 2:** Methodological details and findings of the 7 studies on TL in MS patients.

| First author (year) | Type of specimen | Sampling strategy | Method for TL measurement | Variables considered as confounders | TL in MS patients <sup>a</sup> | TL in controls <sup>a</sup> | Association of TL with MS diagnosis | Other associations with TL |
| --- | --- | --- | --- | --- | --- | --- | --- | --- |
| Hug (2003) [50] | peripheral blood CD4+ and CD8+ T cells | cross-sectional | TRF analysis | age | CD4+: 10.1 kb<br>CD8+: 9.2 kb | CD4+: 8.8 kb<br>CD8+: 9.4 kb | - | age ↓ |
| Guan (2015) [51] | peripheral blood leukocytes | cross-sectional | TRF analysis | - | RRMS: 9.5 ± 0.8 kb<br>SPMS: 9.3 ± 0.9 kb<br>PPMS: 6.7 ± 0.7 kb | 9.1 ± 2.5 kb | PPMS ↓ | age ↓ <sup>b</sup> |
| Guan (2018) [52] | peripheral blood leukocytes | cross-sectional and longitudinal, 3-month follow-up | TRF analysis | - | 6.9 ± 1.0 kb | 9.1 ± 1.4 kb | MS ↓ | - |
| Redondo (2018) [53] | bone marrow cells (MSCs) | cross-sectional | TRF analysis | age, passage number <sup>c</sup> | not specified | not specified | - | age ↓, passage number ↓ <sup>c,d</sup> |
| Krysko (2019) [54] | peripheral blood leukocytes | cross-sectional and longitudinal, 10-year follow-up | singleplex qPCR | age, sex, disease duration | 0.97 ± 0.18 (5.6 kb) | n.c. | n.c. | age ↓, EDSS ↓, disease duration ↓, low brain volume ↓, relapse rate ↓ <sup>e</sup> |
| Habib (2020) [55] | peripheral blood leukocytes | cross-sectional | multiplex qPCR | age, sex | RRMS: 0.76 ± 0.22<br>SPMS: 0.67 ± 0.21<br>PPMS: 0.67 ± 0.27 | 0.94 ± 0.25 | RRMS ↓, SPMS ↓, PPMS ↓ | age ↓, EDSS ↓, passage number ↓ <sup>c</sup> |
| Hecker (2020) [56] | peripheral blood leukocytes | cross-sectional and longitudinal, >10-year follow-up | multiplex qPCR | age, sex | RRMS: 0.92 ± 0.19<br>PPMS: 1.16 ± 0.35 | 1.12 ± 0.35 | RRMS ↓ | age ↓, transition to SPMS ↓ <sup>e</sup> |

In all but one study, average TL were examined in blood cell populations. In 4 studies, the sampling was carried out at a single time point, whereas in the other 3 studies, samples were collected in a longitudinal manner at least for a subgroup of patients. TRF analysis by Southern blotting or qPCR assays [37,38] were employed for TL quantification. The mean TL (± standard deviation, if available) is given for each study group. The statistical analyses usually included adjustments for confounding factors that were suspected to contribute to variation in the data. In 4 out of 6 studies that compared cases and controls, significantly shorter telomeres were observed in at least one MS subgroup. Other associations that were reported in the studies are presented in the rightmost column.
↓ = variable or group was found to be associated with significantly shorter telomeres
<sup>a</sup> = depending on the measurement method used, the TL group averages from the cross-sectional analyses are given in kb (TRF) or in relative values (qPCR)
<sup>b</sup> = significance was reached only for controls and male SPMS patients
<sup>c</sup> = referring to in vitro experiments with patient-derived cells
<sup>d</sup> = decrease of TL with increasing passage number reached statistical significance only for MSCs from MS patients but not for control MSCs
<sup>e</sup> = based on an analysis restricted to the subgroup of RRMS patients
EDSS = Expanded Disability Status Scale; kb = kilobases; MS = multiple sclerosis; MSCs = mesenchymal stromal cells; n.c. = no control group; PPMS = primary progressive MS; qPCR = quantitative polymerase chain reaction; RRMS = relapsing-remitting MS; SPMS = secondary progressive MS; TL = telomere length; TRF = telomere restriction fragment

In all 7 studies, the average length of telomeres over all cells of a cell population and all 92 telomeres per cell has been evaluated. So far, no study investigated TL heterogeneity in single cells and telomeres of single chromosome arms in MS patient samples. In 4 of the 7 studies [50-53], TL were determined by non-radioactive TRF analysis, with only minor differences in the applied protocol: Following electrophoretic separation of digested DNA and Southern blotting, DNA fragments were hybridized to digoxigenin-labeled telomere-specific probes, incubated with anti-digoxigenin antibodies conjugated to alkaline phosphatase, and detected on the blot using chemiluminescent enzyme substrates. In the other 3 studies, qPCR assays were employed: Krysko et al. [54] used the qPCR method by Cawthon from 2002 [37], while Habib et al. [55] and Hecker et al. [56] used the adapted mmqPCR assay by Cawthon from 2009 [38]. In both qPCR-based approaches, relative TL are measured as T/S ratios.

In the studies that have been published so far, no significant TL differences between women and men were described. However, with one exception, all studies reported a clear negative association of chronological age and TL as a marker of biological age, as expected. To consider such data dependencies in the groups to be compared, control for confounding was implemented at the level of the statistical analysis in 5 of the 7 studies (Table 2): Habib et al. [55] and Hecker et al. [56] employed age- and sex-adjusted linear models to assess differences in LTL between patients and controls or associations to clinico-demographic parameters. Redondo et al. considered the independent effects of participant age and passage number in the comparative analysis of TL in cultured MSCs from MS patients and controls [53]. Hug et al. included age as covariate affecting TL in CD4+ and CD8+ T cells [50]. Finally, Krysko et al. used regression models adjusted for age, sex and disease duration [54]. They also evaluated smoking, BMI, type of MS treatment and HLA-DRB1*15:01 carrier status as potential confounders. However, as these variables were not associated with LTL in the study cohort, they were not retained in the final models. On the other hand, one should be aware that the possibility of residual or unmeasured confounding cannot be excluded in general.

Collectively, the present studies provide compelling evidence that the telomeres of immune cells in the peripheral blood are typically shorter in patients with MS. Significantly shorter LTL in at least one MS subtype were consistently found in 4 of the studies (Table 2). In the studies by Guan et al., the measured TL were on average 26% and 24% shorter in the PPMS subgroup [51] and in the general MS group [52], respectively, as compared to the controls. In line with this, Habib et al. found significantly shorter telomeres in each MS subgroup (RRMS: 19%,

SPMS: 29% and PPMS: 29% shorter) [55], and, finally, Hecker et al. observed 18% shorter LTL in the RRMS patient cohort than in the control cohort [56]. A meta-analysis of these 4 studies substantiated that LTL are significantly shorter in MS patients as compared to healthy controls (overall SMD=−0.66, *p*=0.003). Egger’s test revealed no funnel plot asymmetry (*p*=0.771), indicating no major publication bias (Figure 3). However, there was considerable amount of heterogeneity in the study findings (Q test *p*<0.001), and no discernible differences could be detected between subtypes of MS. The other 3 studies could not be included in the meta-analysis due to insufficient data available. In the first study by Hug et al., no apparent differences in T cell TL were reported between cases and controls [50]. Redondo et al. did also not find significant TL differences, but they recognized an accelerated telomere shortening in MSCs from patients with progressive MS in vitro [53]. The remaining study by Krysko et al. did not include a control group [54].

**Figure 3:**
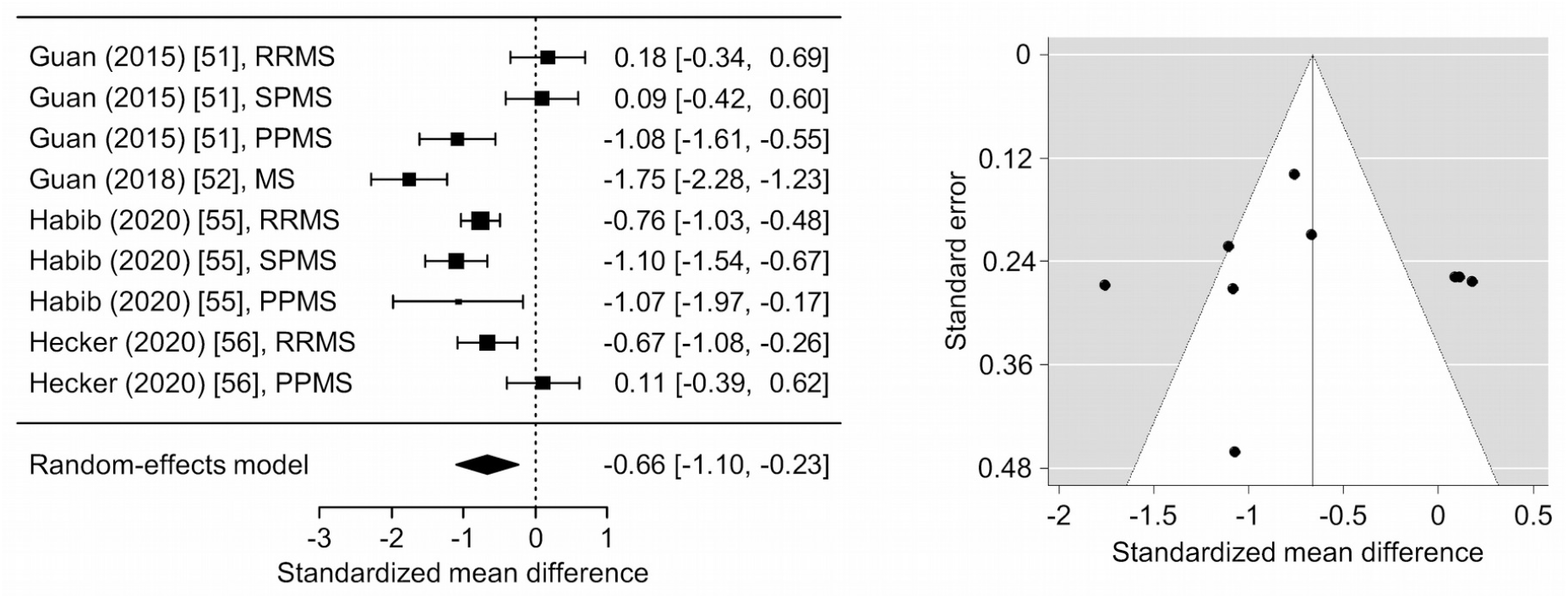
Meta-analysis of the relationship between telomere length and multiple sclerosis. The meta-analysis was based on the results of 4 studies. The forest plot on the left shows the standardized mean differences with 95% confidence intervals for the comparisons of leukocyte telomere lengths (TL) in patients with multiple sclerosis (MS) versus healthy controls in the individual studies and based on a random-effects model. The summary estimate of −0.66 indicated significantly shorter TL in MS (*p*=0.003), and the Cochran’s Q test indicated heterogeneity across the studies (*p*<0.001). The funnel plot on the right showed no apparent asymmetry, and Egger’s regression test detected no evidence of publication bias (*p*=0.771). PPMS = primary progressive MS; RRMS = relapsing-remitting MS; SPMS = secondary progressive MS

The more recent studies in the field also suggest that LTL as a biomarker reflects clinical heterogeneity and that it may be useful to predict disease progression. Krysko et al. [54] found that shorter telomeres are associated with longer disease duration, lower brain volume measures in MRI and higher degrees of disability as measured by the EDSS [4]. A mediation analysis indicated that LTL accounted for 15.1% of the effect of chronological age on disability in their data [54]. The negative relationship between EDSS and TL was also seen by Habib et al., although statistical significance was not achieved when the analysis was adjusted for age [55]. In the longitudinal analyses by Krysko et al., baseline LTL was also weakly associated with EDSS score worsening and cortical gray matter volume decline over time. Moreover, in the subgroup of RRMS patients, a lower LTL was associated with a significantly higher relapse rate in the follow-up [54]. In similar analyses by Hecker et al., RRMS patients with relatively low baseline LTL had nominally higher EDSS scores over 10 years and a significantly higher probability of transitioning to SPMS [56]. Therefore, despite differences in the findings by the different research groups, MS patients with longer telomeres tend to have a more favorable course of the disease.

## 4. Discussion

This is the first systematic review on studies that investigated TL in patients with MS. The analysis of TL provides insights into the proliferation history of cells, as telomeres usually shorten with every DNA replication. TL shortening is thus related to normal aging, but it is also modulated by several environmental influences. In recent years, there has been increased research interest on how the course of MS might be affected by biological aging and the implicated immunological changes. As highlighted in our review, the studies in the current literature show that MS patients typically have shorter telomeres in blood cells as compared to controls. Moreover, it has been argued that the measurement of TL may serve as a potential biomarker for assessing and predicting clinical phenotypes of MS.

The evidence of an association of relatively short TL and MS is convincing despite the fact that only few studies (n=7) with small sample sizes (n≤138 patients for studies comparing against controls) and heterogeneous study populations (e.g., with regard to age, sex and ancestry) have been published so far. Interindividual variation in TL is well known to be affected by demographic factors. On average, longer telomeres were found in adult women compared to men [57], and TL are longer among people of African ancestry compared to people of European ancestry [41]. The studies that were included in this review did not describe any sex-specific differences in TL. However, further studies are needed to better understand to which extent environmental and genetic factors, including sex-related differences, may influence TL as well as risk and severity of MS. It is important to note that the patient cohorts in the reviewed studies also differed in terms of clinical characteristics. For instance, different criteria for the diagnosis of MS were applied, as these were constantly refined over the past years [45-47,58]. Moreover, relapsing and progressive forms of MS were differently represented in the studies. All this may explain some inconsistencies in the findings between the research groups. Therefore, future studies on TL in MS may employ larger patient cohorts that are also more homogeneous in terms of disease activity and progression as evaluated by clinical and MRI parameters. One approach might be to incorporate the analysis of blood cell TL in the context of large-scale observational studies or MS clinical trials.

In most of the reviewed studies, peripheral blood samples were used to measure TL in MS patients. The collection of blood samples is rather safe and convenient, and it is also suitable for repeated assessments in longitudinal studies. However, blood is a heterogeneous composition of various cell types, which may provoke confounding effects in the analysis of TL. Studies have shown that TL and the rate of telomere shortening differ between immune cell types [42,59,60]. For instance, the decline in TL with age in lymphocytes is more pronounced than in granulocytes [42]. As the only research group, Hug et al. used distinct circulating cell types, namely CD4+ and CD8+ T cells, to compare TL between MS patients and healthy controls [50]. Further cell type-specific studies on TL in MS are needed. So far, there is also no TL study that used brain tissues or cerebrospinal fluid cells from MS patients. Hence, the role of CNS cell telomeres in the pathobiology of MS remains to be explored. Telomeres of non-blood cell populations have already been investigated in the context of other diseases. For example, significant differences in TL were found in buccal cells and hippocampal brain tissue from patients with AD in comparison to controls [61]. TL is generally positively correlated across human tissue types, but different tissues have different cellular turnover and replacement, and some tissue types have a certain level of telomerase expression in order to maintain TL in the stem cell compartments [41]. For this reason, the analysis of TL in other tissues and body fluids may give some new information on the association of TL and MS.

In 4 of the 7 included studies, the TRF technique was used, and qPCR assays were chosen in the other 3 studies. Both approaches measure the average length of all telomeres in a cell population. Other methods have been developed to provide deeper insights into the distribution of TL and to quantify TL of specific chromosomes. Therefore, in future studies, methods like Q-FISH [62], for determining the TL of each individual chromosome (p or q arm) in a particular cell, or TeSLA [39], for detecting the shortest telomeres in a sample with high sensitivity, should be taken into account. The shortest telomeres, rather than the average TL, were demonstrated to be critical for genome stability and cell viability as they trigger DNA damage responses leading to replicative senescence [63]. Therefore, more advanced methods may help to delineate cause-and-effect relationships of dysfunctional telomeres in onset and progression of MS. Moreover, they allow measuring subtle changes in the abundance of the shortest telomeres over time, which may have implications for disease monitoring.

Besides TL, there are various other markers of biological aging. In 2013, Horvath developed a predictor of age that allows to estimate the DNA methylation (DNAm) age of tissues and cell types [64]. This “epigenetic clock” focuses on cytosine-5 methylations in DNA regions where a cytosine nucleotide is followed by a guanine nucleotide (CpG). DNAm age utilizes information from 353 CpG sites, of which 193 get hypermethylated and 160 get hypomethylated with increasing age [64]. Using this measure, it has been found that men have higher epigenetic aging rates than women in blood, saliva and brain tissue [65]. In 2018, Levine et al. introduced DNAm PhenoAge, an estimate of phenotypic age based on the methylation status at 513 CpGs that strongly correlates with age but also captures variations in morbidity and mortality outcomes [66]. Theodoropoulou et al. recently compared four age acceleration measures, including the epigenetic clock by Horvath and DNAm PhenoAge, using blood samples from MS patients [67]. They showed that the different measures reflect separate pathophysiological aspects of the disease and that MS patients have significantly higher age acceleration than healthy controls when evaluating DNAm PhenoAge in whole blood. However, a single best measure of biological aging does not exist. TL measurements and the diverse proposed epigenetic clocks reflect different molecular hallmarks of aging [68,69], and each may have its own strengths in the analysis of disease-specific mechanisms.

It currently remains unclear whether shortened telomeres in MS are a cause or a consequence of the pathophysiological processes. An accelerated loss of telomeres with age is known to be attributable to systemic oxidative stress and chronic inflammation, which is associated with increased cellular turnover [28,57]. Interestingly, significantly enhanced lipid peroxidation could be detected in all MS subtypes in the studies by Guan et al. [51,52]. Elevated levels of oxidative stress markers in patients with MS are presumably connected to the inflammatory disease processes. Within the CNS, this in turn may promote demyelination and axonal degeneration and render neuronal repair mechanisms less effective. On the other hand, persistent viral infections were shown to affect TL dynamics and to trigger senescence in immune cells [30].

The accumulation of senescent cells that lack proliferative capacity has adverse consequences as these cells occupy cellular niches and release proinflammatory cytokines [70]. Apart from this, it should be noted that chromatin organization and gene regulation are also modulated by telomere shortening even long before the initiation of DNA damage signals [71]. However, additional research efforts are required to better understand the precise interplay between immune responses and aging mechanisms in MS.

Targeting aging-related mechanisms could be an integral part of future patient care. Micronutrients, such as vitamins and minerals, can modulate oxidative stress and chronic inflammation and thereby affect TL [72]. For instance, higher serum vitamin D concentrations were shown to be associated with longer LTL [73]. Several interventional studies have investigated the effects of vitamin D supplementation in people with MS, but firm conclusions on the clinical benefits cannot yet be drawn [74]. The use of vitamin E and other antioxidants in MS patients is still unclear due to a lack of sufficiently powered studies [75]. Besides, senolytic drugs have been discussed to improve chronic diseases and functional deficits by enhancing immune-mediated clearance of senescent cells [76,77]. Studies with mice showed that removal of senescent cells can prevent or delay tissue dysfunction [78]. Therefore, several senolytics are now tested in clinical trials for treating diverse conditions such as frailty and AD [77], and the administration of senolytics has also been suggested as a potential strategy for delaying progression of MS [79]. The therapeutic modulation of telomerase activity is another approach that is currently being pursued mainly in cancer trials [80]. In the reviewed studies, most MS patients received a basic disease-modifying treatment (DMT), which was not found to be related to TL [54,56]. However, several highly efficacious immune cell depletion therapies have been approved for MS in recent years [81]. Following depletion, the repopulation kinetics vary between different cell types (e.g., B cells repopulate much faster than T cells). These therapies thus have a sustained impact on the immune system. The induced long-term immunological changes probably correlate with changes in TL of circulating lymphocytes. However, the possible relevance of TL dynamics in response to specific MS therapies has not yet been investigated.

To conclude, there is an increased number of studies on the role of telomere attrition in neurological disorders. Here, we identified and reviewed 7 studies that investigated TL in the context of MS. Despite differences in study design and methodology, the findings of the studies overall point to shorter telomeres in blood cells of MS patients in comparison to controls, which was supported by our meta-analysis. Shorter TL were also associated with greater disability and brain atrophy as well as disease progression independent of chronological age. This suggests that biological aging is related to inflammation and neurodegeneration in MS and that the assessment of TL as a biomarker of immunosenescence may provide useful information regarding the individual course of disease. However, further research is needed to better understand the complex relationship between aging and the pathobiology of MS. Subsequent studies may explore the distribution of the shortest telomeres in specific immune cell subtypes and brain cells. This may yield deeper insights into the pathophysiology of MS and allow to develop novel therapeutic strategies targeting aging-related mechanisms.

## Data Availability

Not applicable.

## Abbreviations

AD: Alzheimer’s disease
BMI: body mass index
CIS: clinically isolated syndrome
CNS: central nervous system
CpG: cytosine and guanine separated by a single phosphate group
DMT: disease-modifying treatment
DNA: deoxyribonucleic acid
DNAm: DNA methylation
EDSS: Expanded Disability Status Scale
HLA: human leukocyte antigen
kb: kilobase
LCL: lymphoblastoid cell line
LDL: low-density lipoprotein
LTL: leukocyte telomere length
MHC: major histocompatibility complex
mmqPCR: monochrome multiplex qPCR
MRI: magnetic resonance imaging
MS: multiple sclerosis
MSC: mesenchymal stromal cell
n: number
n.c.: no control group
n.i.: no information
P: passage number of in vitro expansion
PCR: polymerase chain reaction
PPMS: primary progressive multiple sclerosis
PRISMA: Preferred Reporting Items for Systematic Reviews and Meta-Analyses
Q-FISH: quantitative fluorescence in situ hybridization
qPCR: quantitative PCR
RRMS: relapsing-remitting multiple sclerosis
S: single-copy gene signal
SMD: standardized mean difference
SPMS: secondary progressive multiple sclerosis
T: telomere signal
TERC: telomerase RNA component
TERT: telomerase reverse trancriptase
TeSLA: telomere shortest length assay
TL: telomere length
TRF: terminal restriction fragment
U-STELA: universal single telomere length analysis
vs.: versus

## Authors’ contributions

JB performed the literature search, prepared all tables and drafted the manuscript. MH critically revised the manuscript for importent intellectual content and conducted the meta-analysis. BF contributed to the interpretation of published material and the writing of the manuscript. UKZ conceptualized and supervised the research. All authors commented on previous versions of the manuscript. All authors read and approved the final manuscript.

## Acknowledgments

None to declare.

---

**Supplementary Table 1:**
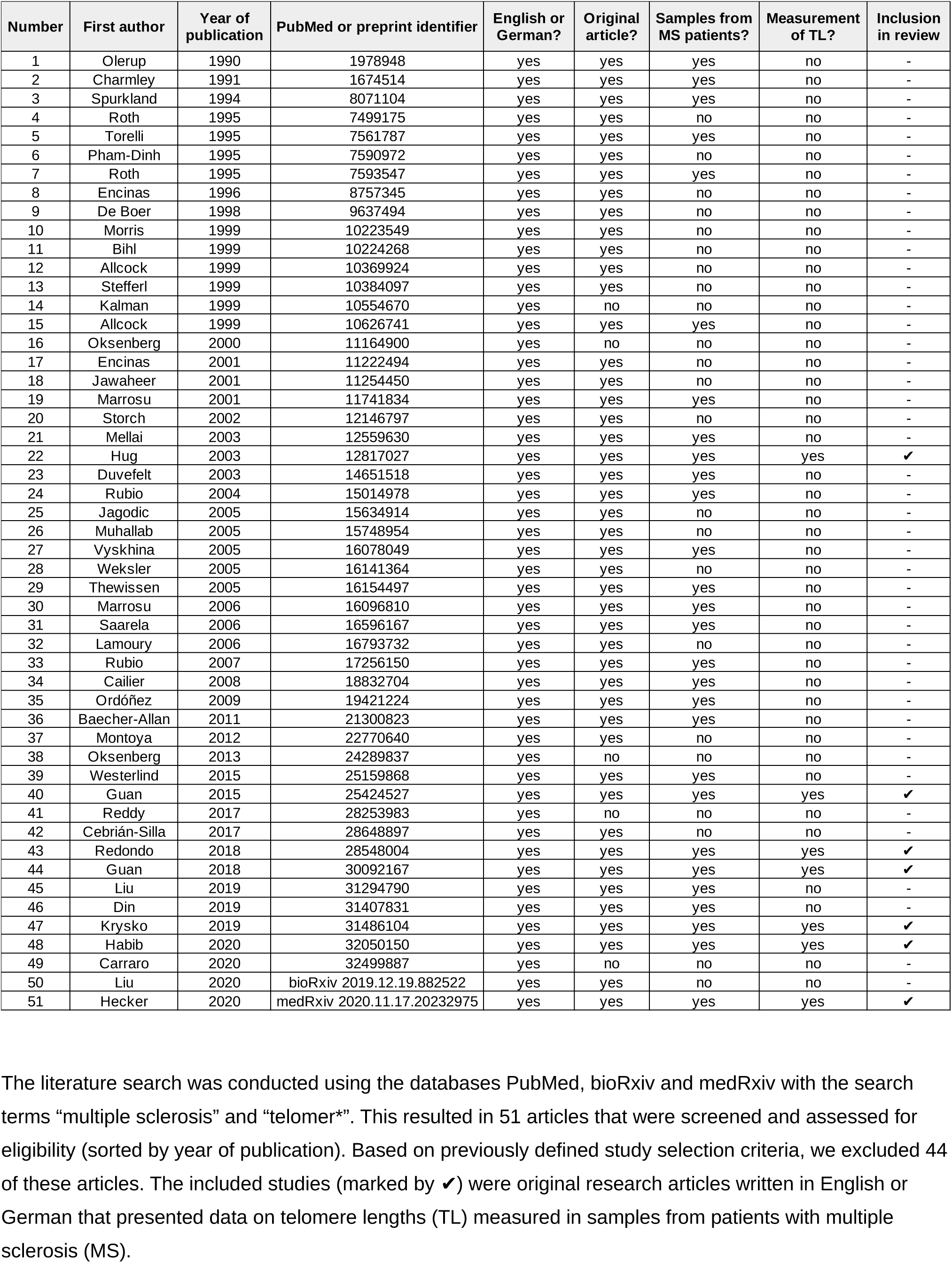
Full list of studies assessed for inclusion in this systematic review.

